# Two decades of molecular surveillance in Kenya reveal shifting *Plasmodium falciparum* drug resistance mutations linked to frontline drug changes

**DOI:** 10.1101/2025.07.15.25331603

**Authors:** George A. Tollefson, Benjamin H. Opot, Titus Kipkemboi Maina, Dennis W. Juma, Raphael O. Okoth, Jackline A. Juma, Gladys Chemwor, Edwin W. Mwakio, Abebe A. Fola, John M. Onge’cha, Bernhards R. Ogutu, Rebecca M. Crudale, Jeffrey A. Bailey, Hoseah M. Akala

**Affiliations:** Department of Pathology and Laboratory Medicine, Brown University, Providence, RI, USA; Center for Computational Molecular Biology, Brown University, Providence, RI, USA; Kenya Medical Research Institute/Walter Reed Army Institute of Research - Africa, Kisumu, Kenya; Kenya Medical Research Institute, Kisumu and Nairobi, Kenya

## Abstract

Malaria control has faced repeated challenges due to the emergence and spread of antimalarial resistance. In this study, we analyzed temporal trends in *Plasmodium falciparum* drug resistance mutations using molecular inversion probes in 642 samples collected in nine sites in Kenya from 1998 to 2021, spanning multiple changes in frontline antimalarial therapies. Following the adoption of artemether-lumefantrine (AL) in 2006, we observed a rapid expansion of MDR1 N86N, Y184F, and D1246D alleles associated with reduced lumefantrine susceptibility, alongside an accelerated decline in chloroquine resistance mutations CRT K76T and A220S. Sulphadoxine-pyrimethamine (SP) resistance mutations, including DHFR N51I, C59R, S108N and DHPS A437G, K540E, rapidly reached fixation following SP introduction in 1998 and persisted through 2021. The high-level pyrimethamine resistance mutation DHFR I164L emerged and expanded after 2012. Further, we identified mutations associated with reduced artemisinin efficacy in the *upb1*, *ap2mu*, and *atp6* genes. Although no validated Kelch 13 (K13) mutations linked to artemisinin partial resistance were detected, we observed the emergence of several previously unreported K13 propeller domain missense mutations, including C473F and A569S, after the adoption of artemisinin-based combination therapies. These findings demonstrate the rapid and dynamic evolution of drug resistance in response to shifting anti-malarial drug pressures and underscore the need for sustained genomic surveillance in malaria endemic regions to inform adaptive treatment strategies. The persistence of SP resistance and the emergence of markers associated with reduced susceptibility to artemisinin and partner drugs raise concerns about the long-term efficacy of current malaria therapies for treatment, seasonal malaria chemoprevention, and intermittent preventive interventions.

## Background

In 2023, 263 million malaria cases and 597,000 deaths attributed to malaria were recorded globally with roughly 3.3 million cases occurring in Kenya.^1^ Global incidence of malaria cases has risen year over year since 2020, signifying a concerning shift after decades of progress towards elimination. Over the last 50 years, widespread use of every frontline antimalarial has selected for resistance in *P. falciparum*. Today, emerging frontline antimalarial drug resistance represents one of the primary challenges to malaria elimination goals, with artemisinin partial resistance (ART-R) already established in Southeast Asia (SEA) and spreading rapidly in several East African countries.

Antimalarial drug resistance often occurs first in Southeast Asia. Chloroquine (CQ) resistance was first reported in the 1950s, soon after widespread distribution of the drug.^2^ In 1979, the first detection of CQ resistant parasites in sub-Saharan Africa was reported in Kenya, where it is believed resistant parasites spread from SEA. In SEA and South America, widespread resistance to CQ necessitated replacement with sulfadoxine-pyrimethamine (SP) as frontline therapy. Over time, resistant parasites both emerged independently in sub-Saharan Africa (sSA) and as well, spread from SEA leading to regional treatment failures to CQ and SP. In sSA, CQ resistance became widespread by the 1980’s after prolonged usage of the drug as first-line medication while declining efficacy of SP was reported within a decade of its implementation in the late 1990’s. By 2004, the World Health Organization (WHO) recommended replacement of SP as frontline treatment for uncomplicated malaria with artemisinin combination therapies (ACTs) due to evidence of increasing rates of SP treatment failure in Africa. ACTs consist of treatment with a powerful, short-acting, artemisinin-derived drug followed by a long-acting partner drug such as the fluorene drug lumefantrine or a quinolone drug, most commonly amodiaquine (AQ), piperaquine (PPQ), or mefloquine (MQ). The WHO decision to adopt combination therapies was based on the idea that individual antimalarial resistance emergence would be reduced as resistance to two drugs would be unlikely to evolve at once in a single infection.^3^

In Kenya, chloroquine (CQ) was initially used as frontline treatment for uncomplicated malaria. CQ resistance was reported in 1979 and, by 1998, widespread CQ treatment failure necessitated replacement with sulfadoxine-pyrimethamine (SP) as frontline therapy.^4^ The WHO recommended the adoption of ACTs in Kenya in 2004 in response to rising SP resistance, but logistical challenges delayed the arrival of the first ACTs for general use in Kenya until 2006.^5^ Artemether-lumefantrine (AL) was chosen as the frontline combination therapy in 2006 due to its efficacy and safety profile and is still recommended in 2025 while dihydroartemisinin combined with piperaquine (DHA–PPQ) is recommended as second-line treatment. While still adequately efficacious, reduced AL treatment effectiveness was first reported in Kenya in 2011,^6^ and by 2016, validated artemisinin partial resistance-associated K13 mutations were observed in neighboring countries.^7^ In 2024, low frequency validated ART-R associated K13 mutations were detected in Kenyan samples collected in 2021.^8^ While discontinued as frontline treatment, the World Health Organization (WHO) recommends SP for Perennial Malaria Chemoprevention (PMC). SP is used for intermittent preventive treatment in pregnancy (IPTp) in malaria endemic areas in Kenya and, in 2023, the country introduced a new seasonal malaria chemoprevention (SMC) strategy treating children under 5 years of age with SP combined with ACT partner drug AQ in selected malaria transmission regions.

Antimalarial drug resistance is driven by genetic mutations which reduce *Plasmodium spp.* susceptibility to antimalarials. Genomic surveillance serves as an effective early detection strategy for emerging drug resistance before the onset of widespread treatment failure. Surveillance is most frequently performed through targeted sequencing of DNA extracted from dried blood spots collected from symptomatic malaria cases at clinics in endemic regions to detect mutations in genes known to be involved in drug resistance. Monitoring temporal and geospatial trends in prevalence of genomic drug resistance markers allows tracking of parasite adaptation to drug pressure and is critical to informing treatment policy changes.

Mutations in a variety of genes have been shown to be associated with reduced parasite susceptibility to most commonly used antimalarial compounds and many candidate mutations are currently being evaluated by in vitro and in vivo therapeutic efficacy studies. While individual drug resistance-associated mutations may confer reduced parasite susceptibility to drug treatment leading to delayed parasite clearance, acquisition of certain combinations of mutations can lead to a compounded effect, in some cases leading to total treatment failure. Chloroquine resistance is primarily mediated by mutations in *P. falciparum* chloroquine resistance transporter protein (CRT) and has been shown to be modulated by co-occurring mutations in the multidrug resistance protein 1 (MDR1). Both are transporter proteins which modulate intraparasitic drug concentrations. CRT-K76T is the hallmark CQ resistance marker and is generally found in all CQ resistant parasites. Additional CRT mutations including A220S, Q271E, N326D, I356L, and R371I have been shown to enhance the resistance phenotype or compensate for parasite fitness costs conferred by mutant disruption to normal cellular processes. MDR1 mutations N86Y and D1246Y have been shown to be involved in enhancing resistance when combined with CRT-K76T.

Sulfadoxine-pyrimethamine (SP) resistance in African *Plasmodium falciparum* has been primarily driven by early fixation of mutations in the *dhfr* and *dhps* genes following the introduction of SP as frontline treatment, notably the combined DHFR IRN (N51I, C59R, S108N) and DHPS GE (A437G, K540E) haplotypes which reduce pyrimethamine and sulfadoxine efficacy respectively.^9^ In East Africa, the appearance of the DHPS A581G mutation comprising the GEG haplotype (A437G, K540E, A581G) has been associated with complete loss of SP efficacy in certain IPTi and IPTp trials.^10,11^ Although still rare in Africa, the emergent DHFR I164L mutation has been shown to confer high-level resistance to SP and antifolate drug combination dapsone–proguanil when co-occurring with the DHFR IRN haplotype.^12,13^

Combinations of mutant haplotypes in different genes have been shown to confer high levels of resistance leading to clinical treatment failure. Previous studies have reported parasite populations carrying multi-locus haplotypes encoding CRT K76T+N326D+I356L+A220S, MDR1 N86Y, and DHFR double mutant C59R+S108N occur the most frequently in SP treatment failures.^14^

Mutations associated with partial resistance to the artemisinin-containing component of ACTs have been identified in several genes in the *Plasmodium falciparum* genome including *k13* and mdr1.^15,16^ Several nonsynonymous mutations in the sequence encoding the propeller domain of the Kelch 13 protein have been shown to be significantly associated with artemisinin partial resistance (ART-R). To date, 12 artemisinin partial resistance-associated mutations have been validated by WHO criteria.^17^ They include F446I, N458Y, C469Y, M476I, Y493H, R539T, I543T, P553L, R561H, P574L, C580Y, R622I, and A675V. Ten other candidate ART-R associated K13 propeller domain mutations have been identified. Of particular interest are candidates P441L and C469F, whose recent emergence and spread are being closely monitored in East Africa.^18,19^ While the exact mechanism is unknown, K13 propeller domain mutations are believed to mediate partial resistance by disrupting the importation of heme into the parasite food vacuole leading to a reduction in the rate of artemisinin activation within the parasite.^20^ Recently, validated ART-R associated mutations have been detected in Kenya. Molecular surveillance identified artemisinin resistance-associated K13 mutation A675V in Kenya in samples collected in 2021,^8^ as well as C469Y in Western Kenya in samples collected in 2022,^21^ highlighting the urgent need for enhanced surveillance.

Mutations associated with reduced susceptibility to ACT partner drugs have been identified in the *mdr1* and *crt* genes. Spread of certain ACT partner drug resistance-associated MDR1 haplotypes has been observed following the implementation of ACTs. In particular, the MDR1 N86N (wild type), Y184F (mutant), D1246D (wild type) (NFD) and N86Y, Y184Y, D1246Y (YYY) haplotypes, which have been shown to be associated with reduced susceptibility to lumefantrine or amodiaquine respectively by reducing the intracellular drug concentration and activity, have been selected for by AL treatment throughout Africa.^22,23,24^ Further, *mdr1* gene amplifications have been found to be associated with reduced susceptibility to mefloquine, lumefantrine, and artemisinin, while decreased copy number has been associated with increased susceptibility to each drug.^25^ Both CRT and MDR1 protein mutations have been shown to be implicated in drug resistance to CQ, amodiaquine (AQ), and piperaquine (PPQ).^26,27^ The mutant haplotype CVIET across CRT residues 72-76 is the most common mutant haplotype in Africa and confers a high level of CQ resistance.^28^ SVMNT has also been detected in Africa and may confer resistance to ACT partner drug amodiaquine.^29^

Despite ongoing changes to antimalarial policy, there remains a critical gap in long-term population level dynamics of *P. falciparum* drug resistance evolution in Kenya. Continuous, region-specific molecular surveillance is urgently needed to inform effective policy adjustments to preserve frontline and preventative antimalarial treatment efficacy. This study provides a comprehensive analysis of changes in drug resistance marker prevalence over two decades of frontline treatment policy changes, presenting key evidence to guide future malaria control strategies in East Africa.

## Methods

### Sample collection

A total of 719 archived peripheral blood DNA samples were selected for sequencing. Samples were collected between 1998 and 2021 from clinical malaria patients over 6 months of age living nearby 11 collection sites primarily located in Western Kenya representing a variety of transmission settings (**Figure 1A and B**).^30–32^ Collection sites included outpatient departments at hospitals in Nairobi, Kisumu, Alupe, Mumias, Kombewa, Kericho, Kisii, Marigat, Busia, Entasopia, and Isiolo. Samples were collected from patients presenting with symptoms of malaria or who tested positive for malaria via microscopy or malaria rapid diagnostic test. Samples were collected under the approval of the Kenya Medical Research Institute (KEMRI) Scientific and Ethics Review Unit (SERU) and the Walter Reed Army Institute of Research (WRAIR) institutional review boards, under protocol numbers WRAIR #1384, #2454 and KEMRI #1330, #3628: Epidemiology of Malaria and Drug Sensitivity Patterns in Kenya. The number of samples collected in each year and from each location are reported in **Supplementary Table S1**.

**Figure 1.**
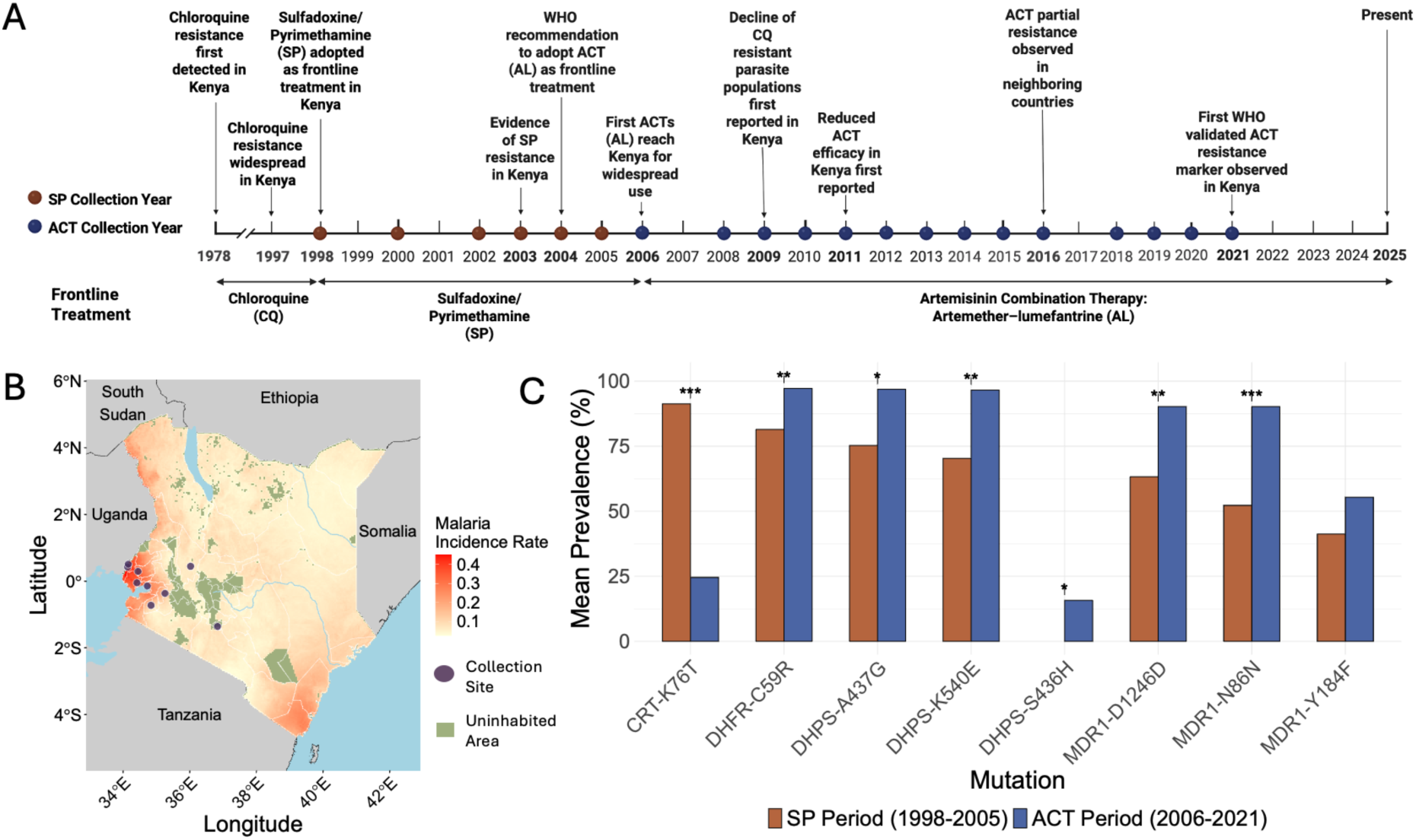
Timeline and locations of sample collection across varying malaria transmission settings spanning changing mutation frequencies in the context of changing frontline antimalarials. A) Timeline of sample collection years and key antimalarial policy and resistance events in Kenya. B) Map of collection sites indicated by blue points overlaid on a geospatial choropleth map of malaria incidence rate (number of newly diagnosed *P. falciparum* cases per 1,000 population averaged over 2000-2021 from the Malaria Atlas Project). C) Mean prevalence of key shifting drug-resistance mutations in *Plasmodium falciparum* among samples from all sites which shifted between SP and ACT treatment eras. Bars represent the mean prevalence (%) of each mutation in the SP era (1998–2005) in brown and ACT era (2006–2021) in blue. Statistical significance was assessed using the Wilcoxon rank-sum test, with p-values adjusted for multiple comparisons using the Holm method. Wilcoxon rank-sum test significance levels are indicated by asterisks (*** = p < 0.001, ** = p < 0.01, * = p < 0.05).

### Extraction of DNA and *Plasmodium* typing PCR

DNA extraction from whole blood was performed using the Qiagen DNA Mini extraction spin protocol (Qiagen, Valencia, CA), following the manufacturer’s guidelines. The DNA was eluted with 150 µl of elution buffer. DNA aliquots were stored long term at -20C as part of batching up for analysis.

The molecular diagnosis of the *Plasmodium* genus and species identification was performed using a quantitative real-time PCR with primers targeting the 18S ribosomal RNA. This procedure involved two reactions. The first reaction detected the presence of the *Plasmodium* genus using a real-time PCR assay on the Applied Biosystems QuantStudio 6 Flex (Thermo Fisher Scientific), as previously described.^33,34^ The second reaction, conducted on the same platform with assay conditions and primers as previously described,^35^ identified the specific *Plasmodium* species present in the infection.^36^ Samples that were positive for *Plasmodium falciparum* with a cycle threshold value of 31 or less were stored at -20C as candidates for analysis.

### MIP capture, sequencing, and analysis

Extracted DNA was captured with molecular inversion probes (MIPs) using the DR2 panel consisting of 814 probes targeting *P. falciparum key* antimalarial drug resistance genes and associated mutations (**Supplementary Table S2**). MIP capture, library preparation, and 150bp paired end sequencing on an Illumina NextSeq550 instrument were conducted as previously described.^37^

Sequence was processed and initially analyzed with the MIPTools pipeline v0.5.0 (https://github.com/bailey-lab/MIPTools), which encapsulates a number of key programs and steps.^37^ In brief, MIPWrangler (https://github.com/bailey-lab/MIPWrangler) was used to stitch paired-end reads, remove sequencing errors using unique molecular identifiers (UMIs), and to identify captured microhaplotypes. Microhaplotypes were aligned to the *P. falciparum* 3D7 reference genome (downloaded 11/08/2018 from https://plasmodb.org/plasmo/app/downloads) using Burrows-Wheeler Aligner BWA-MEM algorithm v0.7.17.^38^ Freebayes 1.3.6 was used to call variants.^39^

Downstream variant call data was utilized to calculate prevalence of known drug resistance-associated mutations in proteins including K13, CRT, MDR1, DHPS, DHFR, AP2MU, UBP1, and ATP6.

We calculated the prevalence of 77 key antimalarial resistance associated mutations (**Supplementary Table S4**) over two decades, focusing on coding mutations associated with resistance to current and previous frontline therapies ACTs, SP, and CQ. Prevalence was defined as *p = m/n x 100* where *m* is the number of samples carrying a variant allele and *n* is the number of samples with a called genotype at the locus. Prevalence of multiple allele haplotypes was calculated similarly where *m* is the number of samples carrying each haplotype and *n* is the number of samples with genotypes called at all haplotype alleles. Haplotypes were exclusively evaluated in samples which were determined to carry a single allele at all loci in each evaluated gene to remove genotypes of mixed infections from haplotype prevalence calculation and visualization. Temporal analyses of mutant prevalence were grouped into multi-year spans to achieve sufficient sample size in each presented time point, accounting for low sample collections in certain individual years.

Individual allele and multiple allele haplotype prevalence was visualized using boxplots, line plots, and stacked barplots using the R package ggplot2 v3.4.0. Shapefiles of Kenyan administrative boundaries and malaria incidence rate data raster files for plotting a map of collection sites across varying transmission settings were downloaded from Malaria Atlas Project (https://github.com/malaria-atlas-project/malariaAtlas). R packages ggplot2 v3.4.0, RColorBrewer v1.1.3, sf v1.0.14, and grid v4.2.2 were used for geospatial plotting.

## Results

### Molecular inversion probe sequencing and analysis

Of 719 samples sequenced, 642 samples (89%) were sequenced with sufficient coverage and quality to include in downstream analysis. Between the two studied time periods (SP Period: 1998-2005, ACT Period 2006-2021), there were 576 samples included in analysis from the ACT period and 66 samples included from the SP period.

### Summary of major changes in antimalarial resistance markers across treatment eras

To provide an overview of key trends in drug resistance, we first summarized the most significant shifts in antimalarial resistance markers across the SP and ACT treatment periods. We observed significant differences in the prevalence of MDR1, DHFR, DHPS, and CRT mutations between samples collected during the years for which SP was designated as frontline antimalarial treatment (1998–2005) compared to those collected when ACTs were used as frontline treatment (2006–2021) (**Figure 1C**).

CRT mutations associated with chloroquine resistance declined after the transition to ACTs. CRT-K76T decreased significantly (Wilcoxon-derived Holm-Bonferroni adjusted p value (padj) = 0.0002), from 94.3% in the SP era to 24.6% in the ACT era. DHFR and DHPS mutations associated with SP resistance reached high prevalence soon after SP introduction and increased in prevalence between the SP and ACT periods. DHFR mutations associated with pyrimethamine resistance remained at high prevalence across both treatment eras. dhfr-C59R increased significantly (padj = 0.046), from 85.6% in the SP era to 97.2% in the ACT era.

DHPS-A437G increased significantly (padj = 0.046), from 76.1% in the SP era to 96.9% in the ACT era. DHPS-K540E increased significantly (padj = 0.046), from 72.0% in the SP era to 96.5% in the ACT era. DHPS-S436H increased significantly (padj = 0.042), from 0% in the SP era to 15.8% in the ACT era. MDR1 mutations associated with lumefantrine resistance increased following ACT implementation. MDR1-N86N increased significantly (padj= 9.51E-06), from 53.3% in the SP era to 90.2% in the ACT era. MDR1-D1246D increased significantly (padj = 0.0001654), from 64.8% in the SP era to 90.2% in the ACT era.

### Lack of prevalent candidate or validated artemisinin partial resistance markers

We detected novel K13 propeller domain missense mutations of unknown significance following the implementation of ACTs. K13 A569S and C473F were each detected in 1 sample in Kisii in 2012 and Marigat in 2020 respectively (**Supplementary Table S3**). A578S, a previously reported polymorphism common throughout sub-Saharan Africa shown to be unassociated with ART-R, was detected at low prevalence in various sites in Western Kenya in 2000, 2008, 2009, 2014, 2015, 2016, 2019, and 2020.

### Increasing prevalence of MDR1 NFD haplotype associated with reduced susceptibility to lumefantrine

Following ACT implementation, prevalence of key MDR1 alleles associated with reduced susceptibility to lumefantrine including N86N wild type, Y184F, and D1246D wild type rapidly increased between the SP (1998–2005) and ACT (2006–2021) eras (**Figure 2A**). N86N wild type increased significantly from 53.3% to 90.2% (Wilcoxon-derived Holm-Bonferroni adjusted p value (padj) = 9.51E-06), and D1246D from 64.8% to 90.2% (padj = 0.0001654), while Y184F increased modestly (42.7% to 55.4%). Haplotype analysis (**Figure 2B**) revealed a transition from the dominant triple mutant YYY haplotype associated with resistance to chloroquine and amodiaquine (50% in 1998–2001) to NFD (71.1% in 2010–2013, 94.6% in 2018–2021), with YYY disappearing by 2014. Two year averaged temporal analysis of key alleles across the collection period (**Figure 2C**) showed that wild type alleles N86N and D1246D declined slowly during the SP era, but then quickly became undetectable in early ACT years, then rapidly increased in prevalence, reaching fixation (100% and 97.7%, respectively) by 2016– 2021. Y184F followed a similar pattern, re-emerging in 2008–2009 and stabilizing at ∼60% from 2014 onward. These findings highlight the replacement of double mutant YYY MDR1 alleles and haplotypes by the NFD haplotype within 6 years following ACT introduction.

**Figure 2.**
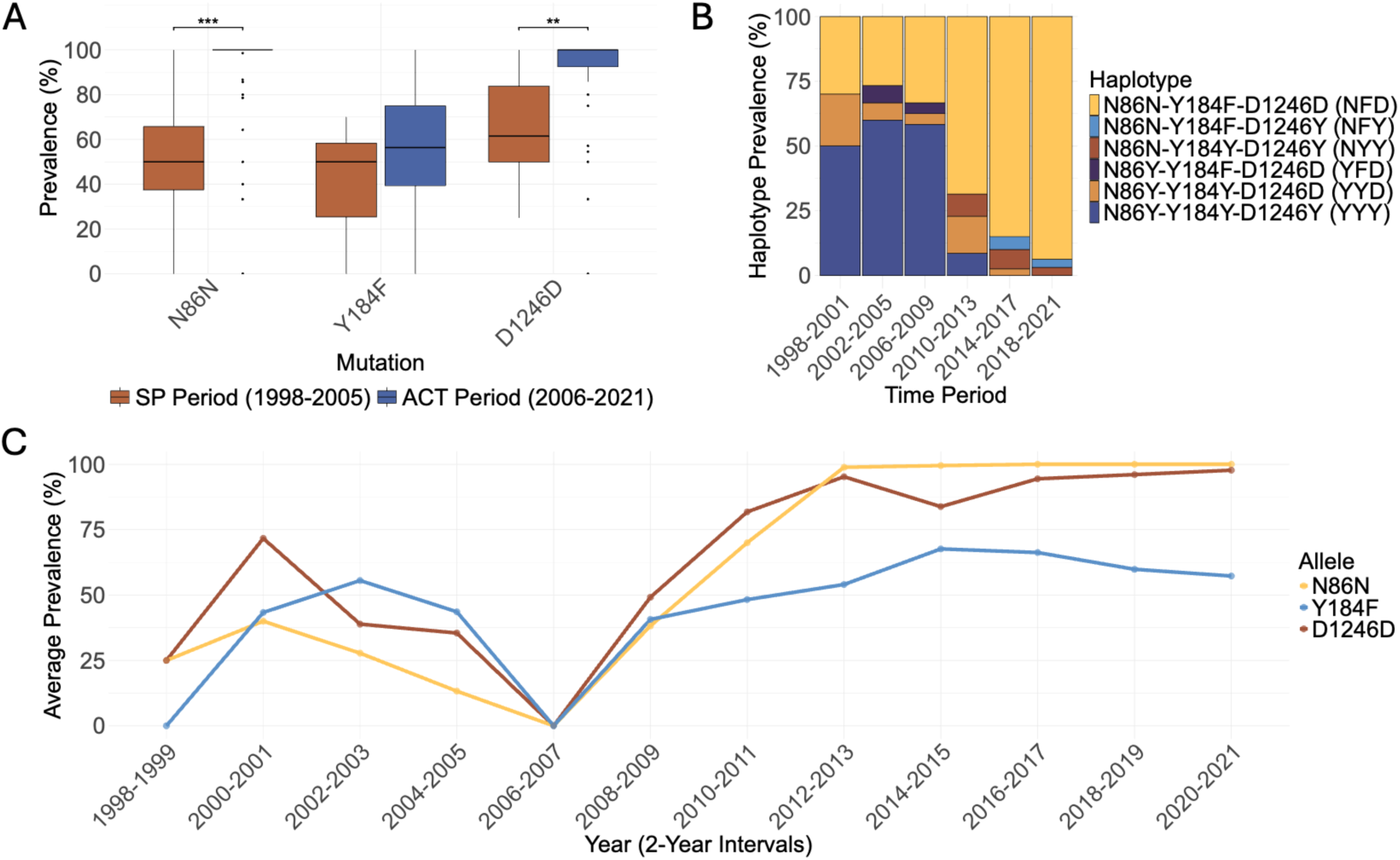
Frequency of key MDR1 alleles associated with reduced susceptibility to lumefantrine. (A) Prevalence of key MDR1 alleles across SP (brown: 1998–2005) and ACT (blue: 2006–2021) treatment periods. Boxplots show prevalence distribution across treatment periods, with Wilcoxon derived Holm-Bonferroni adjusted p values indicated above comparisons where padj< 0.001 = ***, p < 0.01 = **, and p < 0.05 = *. (B) Temporal trends in relative MDR1 haplotype prevalence. C) Temporal trends in individual allele prevalences shown as 2-year averages.

### Rapid reversion to chloroquine sensitivity following ACT implementation

CRT mutations associated with CQ and quinolone-class ACT partner drug (e.g. amodiaquine) resistance remained high after CQ was replaced with SP in 1998 but declined sharply within years following frontline introduction of ACTs in 2006 (**Figure 3A**). Key CRT alleles M74I, K76T, A220S, Q271E, N326S, and R371I all decreased significantly between the tested time periods (SP period: 1998-2005 and ACT period: 2006-2021) with M74I and K76T dropping from 94.3% to 24.6% (padj = 0.0002) and A220S from 94.0% to 27.0% (padj = 0.0003). Q271E, N326S, and R371I followed similar declines, while other evaluated loci remained rare. Haplotype analysis (**Figure 3B**) showed a progressive loss of the hallmark CQ-resistant (CVIET and CVIETS) haplotypes which dominated in 1998–2001 (45% and 15%, respectively) but had nearly disappeared after ACT introduction with WT nearing 98.1% by 2014–2017. We observed a modest resurgence of the CVIETS haplotype after 2018. Temporal analysis of individual key marker prevalences (**Figure 3C**) confirmed this shift, with K76T and M74I persisting at 100% until 2006–2007 before declining steadily to undetectable levels by 2016–2017, alongside similar declines in A220S and Q271E. While N326S showed low-level persistence (5.3% in the ACT era), C72S and V73L remained rare, and D24Y increased modestly (9.6% to 18.6%). We note a steady but continuous increase in prevalence of M74I, K76T, A220S, Q271E, N326S from 2018-2021. These findings highlight the near-complete loss of CRT mutations and the hallmark CVIET haplotype, and the predominance of wild-type alleles despite a small resurgence of parasites carrying the CVIETSS haplotype in later years.

**Figure 3.**
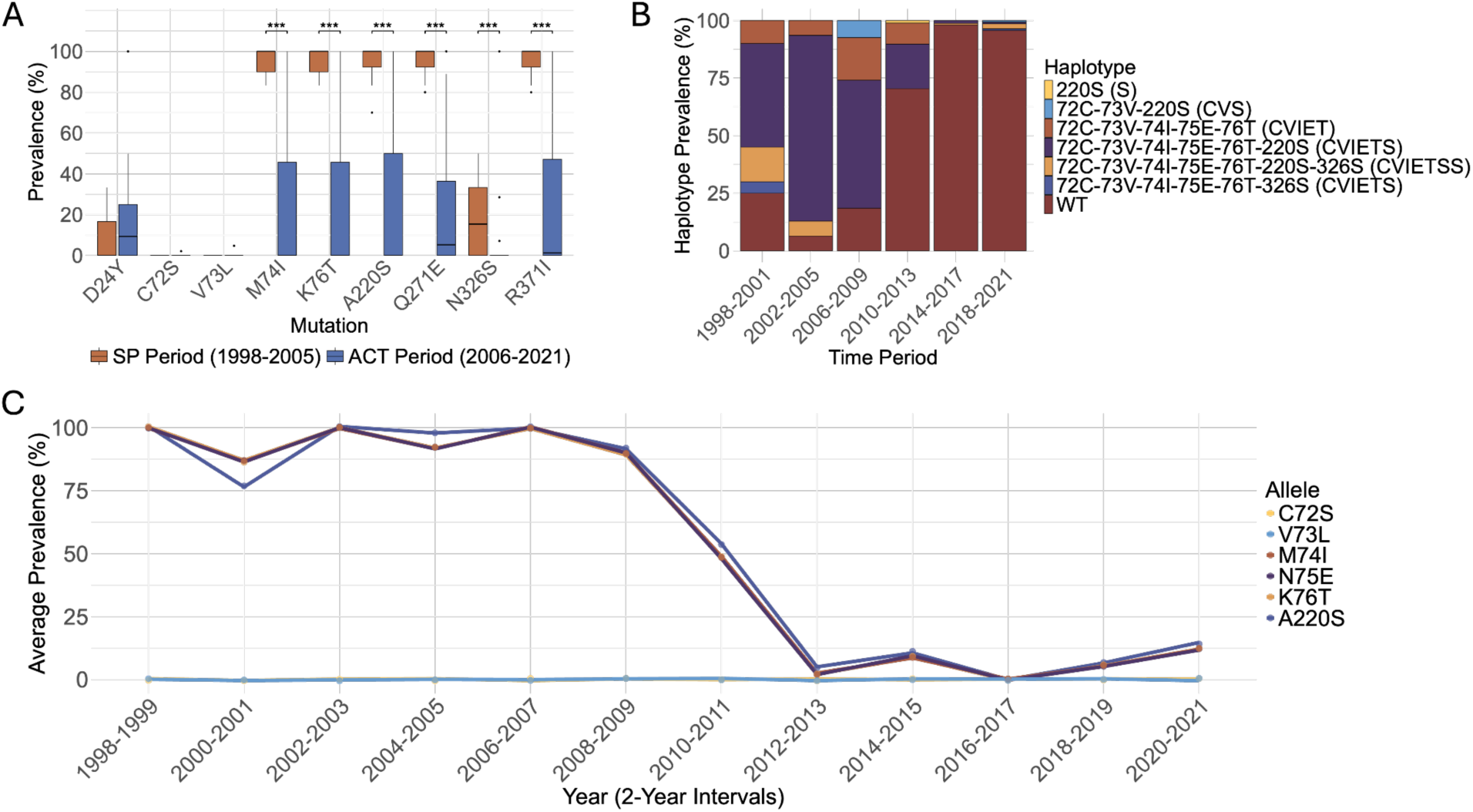
Rapid reversion to chloroquine sensitivity following ACT implementation. (A) Prevalence of key CRT alleles across SP (brown: 1998–2005) and ACT (blue: 2006–2021) treatment periods. Boxplots show prevalence distribution across treatment periods, with Wilcoxon derived Holm adjusted p values indicated above comparisons where p< 0.001 = ***, p < 0.01 = **, and p < 0.05 = *. (B) Temporal trends in relative CRT haplotype prevalence. C) Temporal trends in individual allele prevalences shown as 2-year averages.

### Sustained DHFR and DHPS mutations associated with sulfadoxine-pyrimethamine resistance at near-fixation 20 years after SP implementation

DHFR mutations associated with pyrimethamine resistance remained at high prevalence throughout the study period, with key alleles nearing fixation during the SP era and persisting through the ACT era (**Figure 4A**). N51I remained prevalent in both treatment periods (94.4% in SP, 99.0% in ACT), while C59R increased significantly from 85.6% to 97.2% (padj = 0.046).

**Figure 4.**
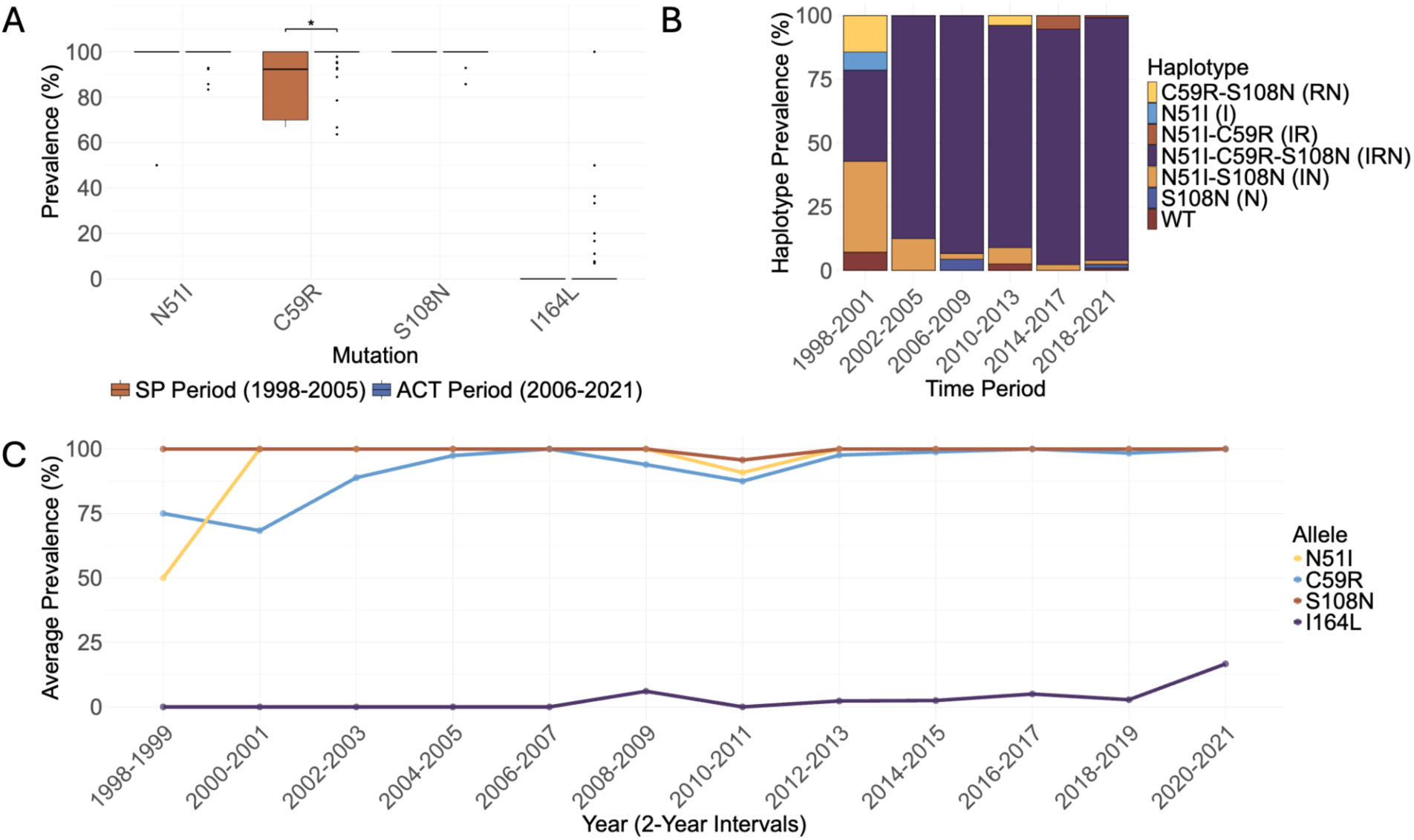
Sustained near-fixation of DHFR mutations associated with decreased susceptibility to pyrimethamine. (A) Prevalence of key DHFR alleles across SP (brown: 1998–2005) and ACT (blue: 2006–2021) treatment periods. Boxplots show prevalence distribution across treatment periods, with Wilcoxon derived Holm adjusted p values indicated above comparisons where p< 0.001 = ***, p < 0.01 = **, and p < 0.05 = *. (B) Temporal trends in relative DHFR haplotype prevalence. C) Temporal trends in individual allele prevalences shown as 2-year averages.

S108N remained near fixation (100% in SP, 99.5% in ACT), and I164L, associated with high-level pyrimethamine resistance, emerged post-ACT introduction at 6.3% prevalence. Haplotype analysis (**Figure 4B**) showed a shift toward IRN (51I-59R-108N), which increased from 35.7% in 1998–2001 to 76.7% by 2002–2005, becoming dominant after ACT introduction (89.4% in 2006–2009) and reaching 91.1% in 2018–2021. Other haplotypes remained at low frequencies. Temporal analysis of allele prevalence (**Figure 4C**) showed N51I reaching fixation by 2000– 2001, with minor fluctuations from 2008-2011 before stabilizing at 100% from 2012 onward.

C59R increased to near fixation from 2006, with small fluctuations, while S108N remained fixed except for a brief dip in 2010–2011. I164L first appeared in 2008–2009 (6.1%), remained low (2.3%–5.0% from 2012–2019), and increased modestly to 16.7% in 2020–2021. These findings highlight the progressive fixation of DHFR mutations and haplotypes linked to pyrimethamine resistance, with sustained high prevalence through the ACT era and a modest rise in I164L from 2017 through 2021.

We observed significant shifts in DHPS mutation prevalence between the SP (1998– 2005) and ACT (2006–2021) eras (**Figure 5A**). S436H, a mutation with uncertain impact on SP resistance but which has previously been reported to be under positive selective pressure in Western Kenya,^40^ increased from 0% to 15.8% (padj = 0.0419), while S436A, present at 17.8% in the SP era, nearly disappeared. A437G and K540E were highly prevalent in both eras, increasing from 76.1% to 96.9% and 72.0% to 96.5% respectively, though not statistically significant. A581G remained rare (1.6% in the ACT era, absent in the frontline SP era).

**Figure 5.**
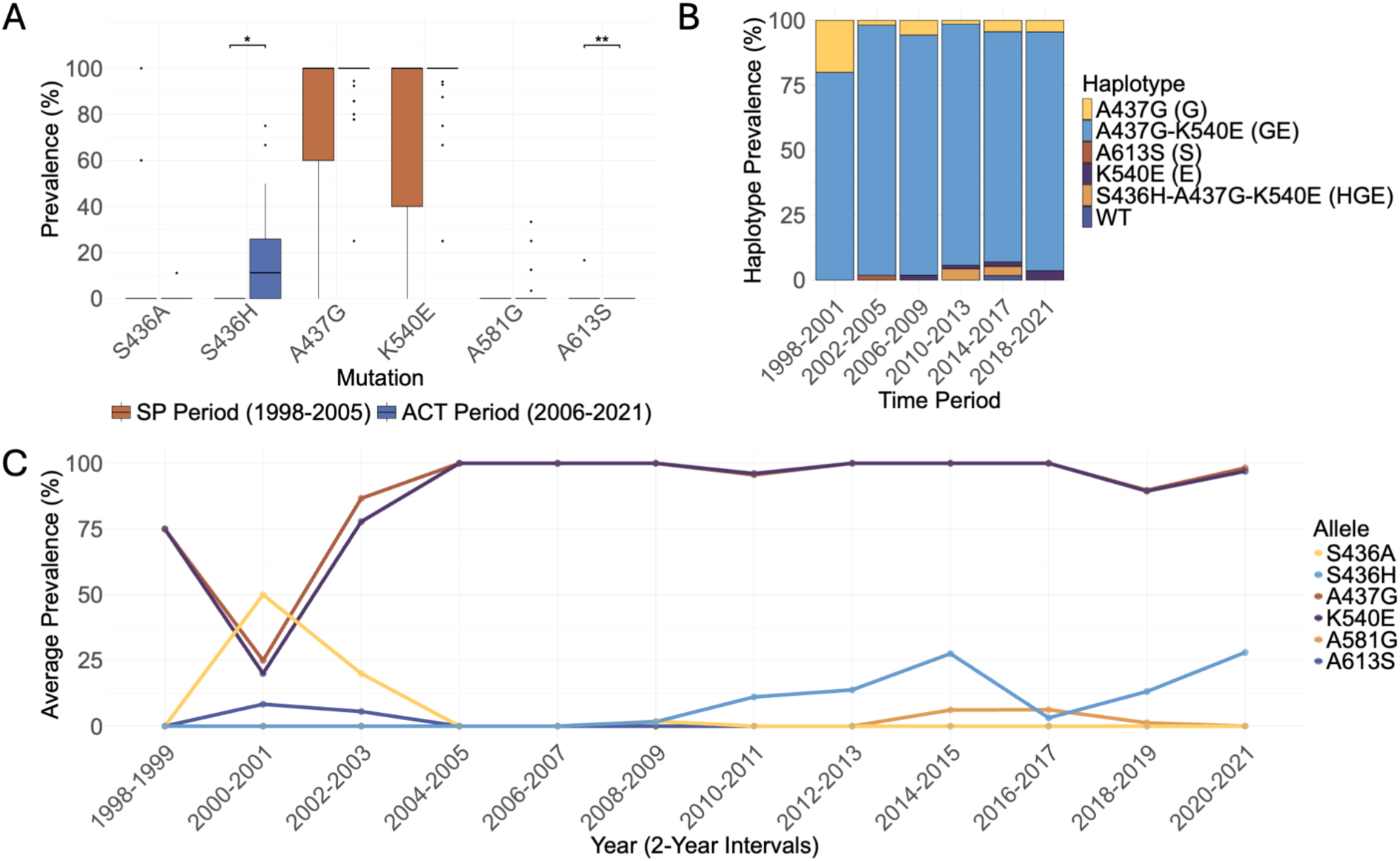
Sustained near-fixation of DHPS mutations associated with decreased susceptibility to sulfadoxine. (A) Prevalence of key DHPS alleles across SP (brown: 1998–2005) and ACT (blue: 2006–2021) treatment periods. Boxplots show prevalence distribution across treatment periods, with Wilcoxon derived Holm adjusted p values indicated above comparisons where p< 0.001 = ***, p < 0.01 = **, and p < 0.05 = *. (B) Temporal trends in relative DHPS haplotype prevalence. C) Temporal trends in individual allele prevalences shown as 2-year averages.

Interestingly, 613S, which has only been commonly observed in west and central Africa, was observed at 16% in 1 of 6 samples tested in Kisumu in both 2000 and 2001. Haplotype analysis (**Figure 5B**) showed a progressive rise in GE (437G-540E), which was already dominant (80%) in 1998–2001 and increased to 92.9% by 2002–2005. After ACT introduction, GE remained high (86.0% in 2006–2009, 83.1% in 2018–2021), with the 437G-alone haplotype persisting at low levels. The HGE haplotype (436H-437G-540E) emerged at 3.9% in 2010–2013 and remained rare. Temporal analysis of allele prevalence (**Figure 5C**) showed A437G and K540E increasing from 75% in 1998–1999 to fixation by 2004–2005, with slight declines in 2018–2019 before rebounding in 2020–2021. S436H emerged in 2008–2009 (1.7%), peaked at 27.6% in 2014–2015, declined, then rebounded to 28.0% in 2020–2021. S436A, detected at 50% in 2000–2001, disappeared by 2004–2005, while A581E first appeared at 6.1% in 2014–2015 but remained rare. These findings indicate the sustained fixation of A437G and K540E, the disappearance of S436A, and the emergence of S436H and A581E at low prevalence towards the end of the study period.

### Temporal trends in mutations associated with artemisinin tolerance

We observed dynamic changes in prevalence of mutations associated with artemisinin tolerance across UBP1, AP2MU, and ATP6 over the study period (**Supplementary Table S5**). UBP1 mutation D1525E rose sharply from near zero at the end of the SP era to 56.3% by 2012– 2013, declined to 8.3% in 2016–2017, before rising to 21.2% in 2020–2021. E1528D prevalence was high initially (66.7% in 1998–1999) before declining to undetectable levels in 2006–2007, ultimately reappearing at low prevalence (11.8–23.3%) from 2012 onward. AP2MU S160N fluctuated, declining from 75% in 1998–1999 to 33.3% in 2002–2003, peaking at 100% in 2006–2007, then decreasing to 9.7% in 2010–2011 before transiently rebounding (38.5% in 2012– 2013) and gradually declining (12.1% in 2020–2021). ATP6 mutations appeared sporadically, with A623E was rare and undetectable after 2008–2009, while E431K fluctuated, peaking at 38.9% in 2002–2003, disappearing in 2006–2007, re-emerging at 30.7% in 2008–2009, peaking again (66.7% in 2016–2017), then declining to 18.8% in 2020–2021. S769N was not detected. These findings highlight the dynamic emergence, disappearance, and resurgence of mutations in UBP1, AP2MU, and ATP6.

## Discussion

Here we sequenced retrospective samples from over a two-decade period that allows us to evaluate long-term temporal trends in drug resistance mutations over the course of changing frontline antimalarials in Kenya. The observed trends broadly align with those observed throughout East Africa. Selective pressures imposed by treatment with ACTs, SP, and other antimalarials continue to drive changes in drug resistance allele prevalence which carry important implications for treatment efficacy and future therapeutic strategies. Recent concerning changes include the persistent rise in the MDR1 NFD haplotype since the implementation of ACTs in Kenya as well as DHFR 164L and the DHPS 581G, both associated with high-level SP resistance which could compromise continued use in IPTp and in seasonal chemoprevention. The observed persistence of hallmark chloroquine resistance CRT mutations at fixation eight years following replacement of CQ with SP and the subsequent sharp decline in CQ resistance marker prevalence following AL implementation demonstrates a strong temporal association with ACT introduction and further strengthens CRT wild type association with decreased lumefantrine sensitivity.

The introduction of artemether-lumefantrine (AL) in 2006 coincided with the rapid shifts in alleles and haplotypes associated with decreased sensitivity to lumefantrine. There was an expansion of the mainly wild type MDR1 N86N-Y184F-D1246D (NFD) haplotype replacing the MDR1 N86Y-Y184Y-D1246Y (YYY) haplotype, indicating reduced lumefantrine susceptibility,^41^ and a return to amodiaquine, piperaquine, and chloroquine sensitivity in Western Kenya. This shift has also been observed in Uganda, Tanzania, and Rwanda.^22,42^ Similarly, temporal trends in CRT haplotypes reflect shifting quinolone drug pressures. The CVIET haplotype which was widespread during the CQ and SP treatment period declined after ACT adoption, with a resurgence of CQ-sensitive wild-type CRT alleles. This suggests SP lacks a negative selective effect on the CQ resistance genotype and that either there was continued chloroquine usage during this period and/or minimal negative selective pressure in the absence of CQ. It supports previous in vitro evidence of the positive selective pressure of lumefantrine for wild type CRT-K76K in clinical isolates from Kenya.^43^ Interestingly, the observed modest resurgence of CRT-K76T after 2018 in our sample collection suggests recent ongoing selection, possibly from PPQ pressure from use of alternative frontline ACT DHA-PPQ in Kenya. Further investigation into relative rates of treatment with AL and DHA-PPQ in Western Kenya in these years may clarify sources of selective pressure.

SP resistance haplotypes have shown remarkable stability and continued evolution. Further increases since withdrawal as frontline therapy in 2006 suggest minimal fitness costs and ongoing drug pressure. The triple DHFR-IRN (I51-R59-N108) and double DHPS GE (G437G-E540E) haplotypes associated with high level SP resistance reached fixation shortly after SP became frontline therapy in 1998 and have persisted despite its withdrawal from routine case management in 2006. Continued SP use in IPTp programs and likely minimal selective disadvantage may sustain this resistance. More concerning is the emergence of quadruple DHFR IRNL (I51I-R59R-N108N-I164L) and triple DHPS GEG (G437G-E540E-G581G), which have been reported in Rwanda and Tanzania and could undermine SP-based IPTp and seasonal chemoprevention if they continue to spread and reach high prevalence.^44,45^

Although we did not detect any validated K13 mutations conferring partial artemisinin resistance (ART-R), our collection ended in 2021 which is the year in which the first K13 validated mutation was reported to have emerged in Kenya. We did identify two novel uncharacterized K13 mutations in 2012 and 2020. We also detected UBP1 mutations D1525E and E1528D and AP2MU mutation S160N which have all been linked to delayed clearance after treatment with ACTs,^46^ as well as ATP6 mutations A623E and E431K which have previously been associated with increased artemether IC_50_.^47^ Given the rapid rise of ART-R K13 mutations in Uganda and Rwanda, these findings suggest that Kenya may be ripe for ART-R spread with ancillary mutations and the undermining of partner drugs. It has previously been shown that pre-existing resistance to partner drugs may facilitate the emergence of ART-R associated K13 mutations.^47^ It is likely recently reported Kenyan *P. falciparum* populations carrying validated ART-R associated mutations, C469Y and A675V,^8,21^ represent spread from their epicenter in Uganda, but this has not been confirmed. Further assessment is needed, and proactive monitoring is essential for understanding their origin and spread, as well as for formulating effective control strategies given their potential to undermine ACTs.

This study has several limitations. Our samples were collected where malaria is currently predominantly found, Western Kenya and may not fully represent the lower malaria transmission settings across the country, where resistance can often emerge. Additionally, our findings do not reflect more recent developments in *P. falciparum* resistance trends given the most recent samples are 2021. We also lack the ability to examine other factors that drive the persistence and expansion of drug-resistant malaria parasites in Kenya such as the non-recommended use of drugs through the private sector, which is a central challenge for the vast majority of studies.

The opposing selection pressures exerted by ACT partner drugs on MDR1 drug resistance haplotypes offer a recognized opportunity for mitigation of resistance development.^48,49^ While AL selects for NFD, thereby reducing lumefantrine susceptibility, DHA-PPQ exerts the opposite pressure, selecting for YY, which restores lumefantrine sensitivity but increases PPQ resistance risk. AQ, used in SMC, also selects against NFD. This suggests that triple-ACTs (TACTs) combining lumefantrine with piperaquine or amodiaquine could help maintain drug efficacy. Given promising preliminary results from Kenya in 2021,^50^ expanded clinical trials evaluating LU-PPQ, LU-AQ, and LU-MQ combinations with artemisinin containing drugs in East Africa should be prioritized.

Continued molecular surveillance is necessary to monitor emerging resistance in Kenya and effort should be made to collect sufficient samples across diverse transmission settings and treatment availability regions. Combining enhanced molecular surveillance with therapeutic efficacy studies (TES) will provide a more comprehensive understanding of drug resistance evolution and inform future malaria control strategies. This work is particularly important considering the recent emergence of validated ART-R associated K13 mutations in Kenya. As ART-R expands regionally and ACT partner drug and preventative treatment resistance trends continue to evolve, enhanced genomic surveillance will be critical for guiding adaptive treatment policies and sustaining malaria control efforts.

## Ethical clearance

This study was conducted under the approval of the Kenya Medical Research Institute (KEMRI), Scientific and Ethics Review Unit (SERU) and Walter Reed Army Institute of Research (WRAIR) institutional review boards, protocol numbers: WRAIR #1384, #2454 and KEMRI #1330, #3628: “Epidemiology of malaria and drug sensitivity patterns in Kenya.”

## Data Availability

Raw sequencing data are available under accession number (PENDING) at the Sequence Read Archive (SRA) (http://www.ncbi.nlm.nih.gov/sra), and the associated BioProject is (PENDING). De-identified data sets produced in this study which were used to make all figures are available as supplementary tables.

We report drug resistance mutation genotype coverage and individual level within-sample allele frequency data (**Supplementary Table S6**) for all called missense SNPs in drug resistance genes targeted by our MIP panel following best reporting standards for molecular surveillance data so that our results are reproducible and may be utilized for future retrospective studies.^51^

## Code availability

Code for data analysis is available on GitHub under (PENDING). Additional software tools used for MIP data analysis are available at https://github.com/bailey-lab/MIPTools.

## Author contributions

- George A. Tollefson: Conceived and designed the study, data collection, data interpretation, and initial drawing and revision of the manuscript.
- Benjamin H. Opot: Molecular analyses, data interpretation, manuscript preparation, and critical review of the manuscript.
- Titus Kipkemboi Maina: Molecular data collection, analyses, interpretation, and review of the manuscript.
- Dennis Juma: Conceived and designed the study, coordinated data collection, supervised laboratory analyses, contributed to data interpretation, and critically reviewed the manuscript.
- Raphael O. Okoth: Molecular data collection, analyses, interpretation, and review of the manuscript.
- Jackline A. Juma: Molecular data collection, analyses, interpretation, and review of the manuscript.
- Gladys Chemwor: Molecular data collection, analyses, interpretation, and review of the manuscript.
- Edwin W. Mwakio: Molecular data collection, analyses, interpretation, and review of the manuscript
- Abebe A. Fola: Data interpretation and review of the manuscript.
- John M. Onge’cha: Coordinated data collection, contributed to data interpretation, and critically reviewed the manuscript.
- Bernhards R. Ogutu: Conceived and designed the study, data interpretation, and critically reviewed the manuscript.
- Rebecca M. Crudale: Molecular data collection, analyses.
- Jeffrey A. Bailey: Conceived and designed the study, coordinated data collection, data interpretation, and critically reviewed the manuscript.
- Hoseah M. Akala: Conceived and designed the study, coordinated data collection, contributed to data interpretation, and critically reviewed the manuscript.

## Acknowledgments

We thank the individuals who participated in this study for providing these invaluable samples and time to generate these findings. We also thank the Ministry of Health Kenya for giving us access to these samples. We also thank all clinical staff at all the participating hospitals for their assistance. We thank the Director, Walter Reed Army Institute of Research-Africa, COL. Lacy Shannon, Director Walter Reed Army Institute of Research - Africa Kisumu field Stations, COL. Gerald Keller, Deputy Director General, Center for Clinical Research, Kenya Medical Research Institute Dr. Linus Ndegwa, Kenya for supporting this study and giving their permission to publish these data. We also thank all clinical staff at all the participating hospitals for their assistance.

## Supplementary Tables

For readers convenience, we have collated Supplementary Tables S1-S5 into a single Excel workbook with each table presented in a separate, clearly labeled tab. Supplementary Table S6 is hosted on Zenodo due to its large file size and is accessible under DOI: 10.5281/zenodo.15865821.

**Supplementary Table 1**. Sample collection summary table summarizing sample counts by collection year and site).

**Supplementary Tables 2**. This set of tables includes **A)** Genomic ranges targeted by probes for MIP capture and sequencing; **B)** Details of all MIP probe sequences; and **C)** Genomic positions of key drug resistance associated mutations targeted by the panel.

**Supplementary Table 3**. Table of sample level incidence of all K13 propeller domain missense mutations with year and location of detection.

**Supplementary Table 4**. Table of prevalence of 77 individual validated antimalarial resistance markers by sample year and location.

**Supplementary Table 5**. Table of prevalence of artemisinin resistance mutations in UBP1, ATP6, and AP2MU by sample year and location.

**Supplementary Table 6**. Individual level within-sample allele frequency data for all DR2 panel missense SNPs detected in one or more samples.

